# Serum PFOS Concentration and Isomer Composition in Relation to Depressive Symptoms in Young Adults

**DOI:** 10.64898/2026.10.14.26365394

**Authors:** Yue Bi, Yakun Liu, Yuanyuan Li, Hui Bu

## Abstract

Total serum perfluorooctane sulfonate (PFOS) concentration and its isomer composition describe different aspects of internal exposure, but their respective associations with depressive symptoms remain uncertain. We examined adults aged 18-25 years in NHANES August 2021-August 2023 using survey-weighted models of the Patient Health Questionnaire-9 (PHQ-9). Total measured PFOS was the sum of linear PFOS and pooled monomethyl-branched PFOS; composition was their log-ratio. The analytic cohort included 169 participants, with 147 complete cases for core adjustment. Median total PFOS was 1.70 ng/mL, the median linear fraction was 72.2%, and the weighted prevalence of PHQ-9 ≥10 was 22.6%. In the joint adjusted model, a doubling of total PFOS at fixed composition corresponded to -0.49 PHQ-9 points (95% CI, -1.65 to 0.68); a doubling of the linear-to-branched ratio at fixed total corresponded to 0.21 points (-1.94 to 2.36). Algebraic rescaling across the reported interquartile ranges yielded differences of -0.58 points (-1.95 to 0.81) for total PFOS and 0.13 points (-1.16 to 1.41) for composition. The inverse continuous-score association weakened with adjustment, whereas the screening odds ratio primarily lost precision. Excluding three observations with an isomer below detection shifted both joint coefficients. The data therefore do not establish an independent composition association or equivalent isomer effects. Experimental evidence supports candidate oxidative, inflammatory, and synaptic pathways, but differential protein binding and elimination prevent a serum composition ratio from being interpreted directly as brain dose or toxicity. Larger longitudinal analyses with isomer-resolved exposure and toxicokinetic measurements are needed to test these pathways.

## 1 Introduction

Depressive symptoms during young adulthood arise within a changing social, behavioral, and biological context.^1^ Environmental biomonitoring can help investigate potentially relevant exposures, but it requires a clear distinction between a circulating chemical measurement, a biologically active tissue dose, and a mental-health outcome. The Patient Health Questionnaire-9 (PHQ-9) summarizes symptoms over the preceding 2 weeks; a score of at least 10 indicates a positive screen rather than a diagnosis of major depressive disorder.^2,3^

Perfluorooctane sulfonate (PFOS) is a persistent per- and polyfluoroalkyl substance (PFAS). Ingestion of contaminated water and food, including contaminated fish, and exposure to contaminated dust can contribute to internal exposure.^4^ Serum PFOS can persist for years, so one measurement integrates aspects of past intake and elimination rather than identifying a recent exposure event.^5^ Its relevance to depressive symptoms is biologically plausible: experimental studies have linked PFOS to neuronal stress signaling, microglial dysfunction, and altered hippocampal synaptic pathways.^6,7,8^ Whether these findings translate to the much lower internal concentrations encountered in a contemporary population sample remains unresolved.

PFOS comprises structural isomers with the same elemental composition but different carbon skeletons. NHANES reports linear PFOS (n-PFOS) and the summed monomethyl-branched component (Sm-PFOS) separately. Their sum is the operational measure of total PFOS used here; the branched result is a pooled analytical measurement rather than a single chemical species.^9,10^ Thus, total concentration asks how much measured PFOS is present, whereas composition asks how that amount is allocated between the linear and pooled branched components. The ratio is a descriptor of the same exposure mixture, not another chemical that enters the body independently.

These quantities may have different biological meanings. Linear and branched PFOS differ in albumin binding, urinary elimination, and transfer across some biological barriers.^11,12,13^ Human elimination studies also show that kinetic differences depend on the particular branched isomer and study population.^14,15^ A higher serum linear fraction may consequently reflect source composition, differential retention, or exposure timing. It does not by itself establish a larger unbound fraction, greater brain penetration, or higher neurotoxic potency. Distinguishing toxicokinetics, which determines internal distribution, from toxicodynamics, which determines the response at a given target-tissue dose, is necessary for interpreting an isomer association.

Previous NHANES analyses have reported both inverse and positive PFAS-depressive-symptom associations under different exposure definitions and model specifications.^16,17^ Analyses of separate component concentrations do not directly answer whether relative isomer composition matters when total PFOS is held constant. This conditional comparison is chemically informative because increasing the linear share at a fixed total necessarily reduces the measured branched share. Isomer-resolved NHANES data have been available since 2013–2014; the present contribution is the explicit separation and interpretation of these two exposure dimensions in a recent young-adult sample.^10^

We examined total measured PFOS and the linear-to-branched ratio jointly in adults aged 18-25 years in NHANES August 2021-August 2023. The primary outcome was continuous PHQ-9 score, with PHQ-9 ≥10 as a secondary screening outcome. We interpreted model coefficients both per doubling and over reference contrasts based on the reported interquartile ranges, evaluated sensitivity to outcome definition and detection status, and considered what the observed associations can and cannot reveal about candidate mechanisms. The analysis estimates cross-sectional associations; it does not test neural mediation or establish that PFOS causes depression.

## 2 Methods

### 2.1 Study design and analytic population

This cross-sectional study used public-use data from NHANES August 2021-August 2023, a stratified, clustered, multistage probability survey of the civilian, noninstitutionalized U.S. population.^18^ Adults aged 18-25 years at screening were selected using RIDAGEYR. The public depression-screener file includes participants aged 18 years and older; responses for those aged 12-17 years are available only through restricted access.^3^

PFAS_L, DPQ_L, DEMO_L, BMX_L, and COT_L were linked by respondent sequence number (SEQN). Eligibility required both PFOS measurements, a positive PFAS subsample weight, complete responses to the nine PHQ-9 symptom items, and the survey design variables. Of 11,933 survey participants, 847 were aged 18-25 years, 194 had valid weighted PFAS measurements, and 169 had a complete PHQ-9 score. Core-adjusted analyses included 147 participants with complete covariates (**Figure 1**). The difference between the 847 age-eligible participants and the 194 with valid PFAS data includes laboratory subsampling and specimen or data availability; it should not be interpreted as ordinary item nonresponse.

**Figure 1.**
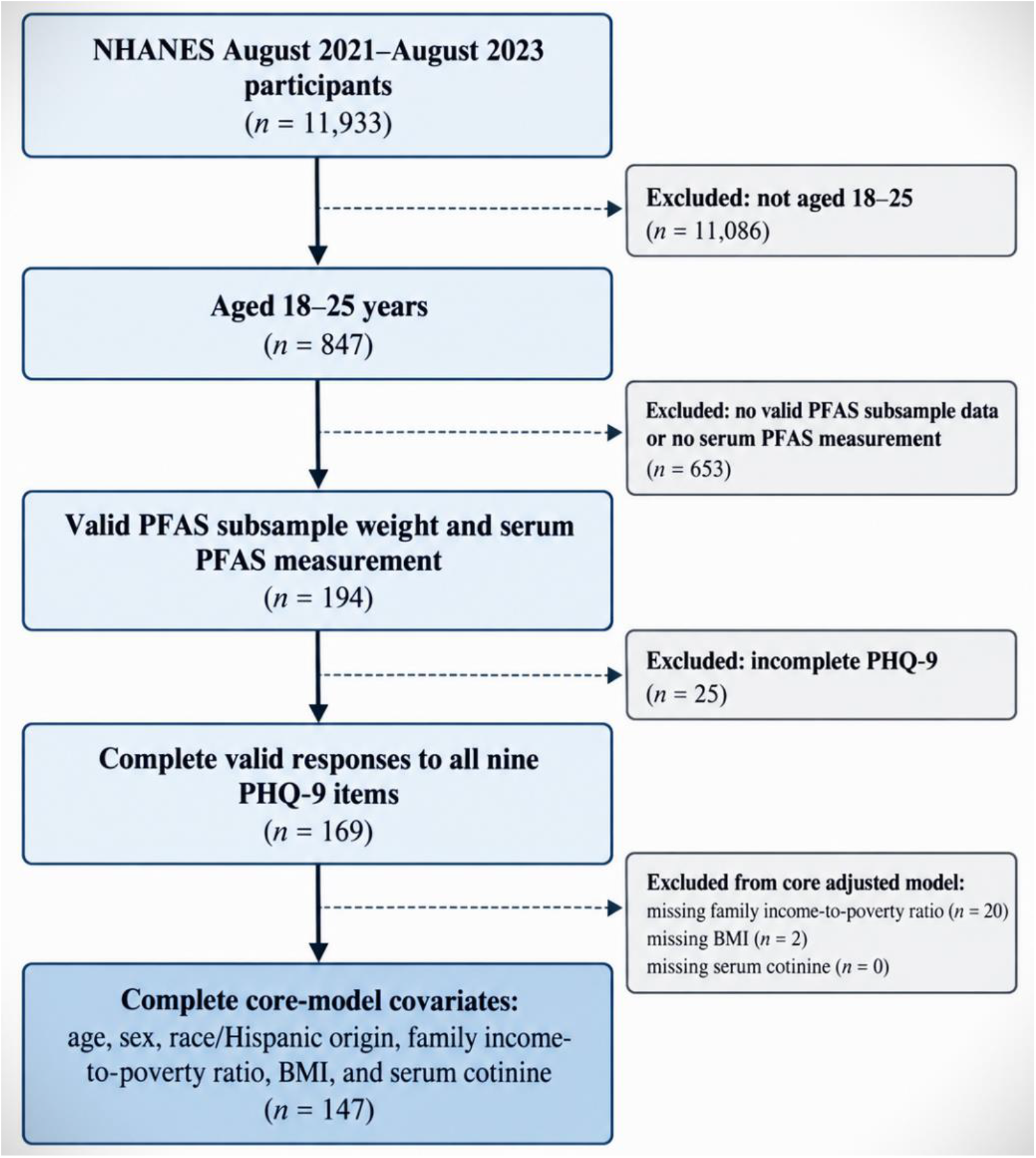
Selection of the analytic sample. NHANES August 2021-August 2023 included 11,933 participants, of whom 847 were aged 18–25 years. Of these, 194 had valid weighted PFAS data, 169 had all nine PHQ-9 symptom responses, and 147 had complete core covariates. The 653 participants without valid PFAS data include those outside the laboratory subsample and those without usable specimen or measurement data. Counts are unweighted. Core covariates are age, sex, race and Hispanic origin, income-to-poverty ratio, BMI, and serum cotinine.

NHANES procedures were approved by the National Center for Health Statistics Ethics Review Board, and participants provided consent.^18^ This analysis used de-identified public-use records and involved no participant contact.

### 2.2 PFOS measurements and exposure contrasts

Serum PFAS were quantified by online solid-phase extraction coupled with high-performance liquid chromatography and negative-ion tandem mass spectrometry.^9^ Linear PFOS was obtained from LBXNFOS and summed monomethyl-branched PFOS from LBXMFOS. The limit of detection (LOD) was 0.10 ng/mL for each component. NHANES-assigned values below the LOD, equal to LOD/√2 subject to the precision of the released data, were retained. Detection status was identified from LBDNFOSL and LBDMFOSL.

Let L denote n-PFOS and B denote Sm-PFOS, both in ng/mL. Total measured PFOS (T), linear fraction (f), and the principal composition metric (R) were defined as follows:

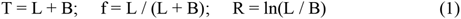

The term total measured PFOS refers to the sum of these two reported components, rather than an exhaustive quantification of every possible PFOS isomer. Magnitude entered regression models as X = log₂(T). Composition entered as R, with estimates rescaled for presentation to a doubling of L/B. Thus, if βR is the coefficient for the natural-log ratio, the reported difference per doubling of the ratio is βR ln(2).

The two coefficients describe different conditional comparisons (**Figure 2**). Doubling T at fixed composition doubles L and B together. Doubling L/B at fixed T increases the linear share while decreasing the branched share. For example, at a hypothetical T of 1.70 ng/mL, changing f from 70% to 80% changes L from 1.19 to 1.36 ng/mL and B from 0.51 to 0.34 ng/mL. This is a constructed contrast, not a comparison of identified participants. At f = 72.2%, doubling L/B increases f to approximately 83.9%, an 11.7-percentage-point change. The composition coefficient therefore cannot be interpreted as the effect of adding linear PFOS while leaving branched PFOS unchanged.

**Figure 2.**
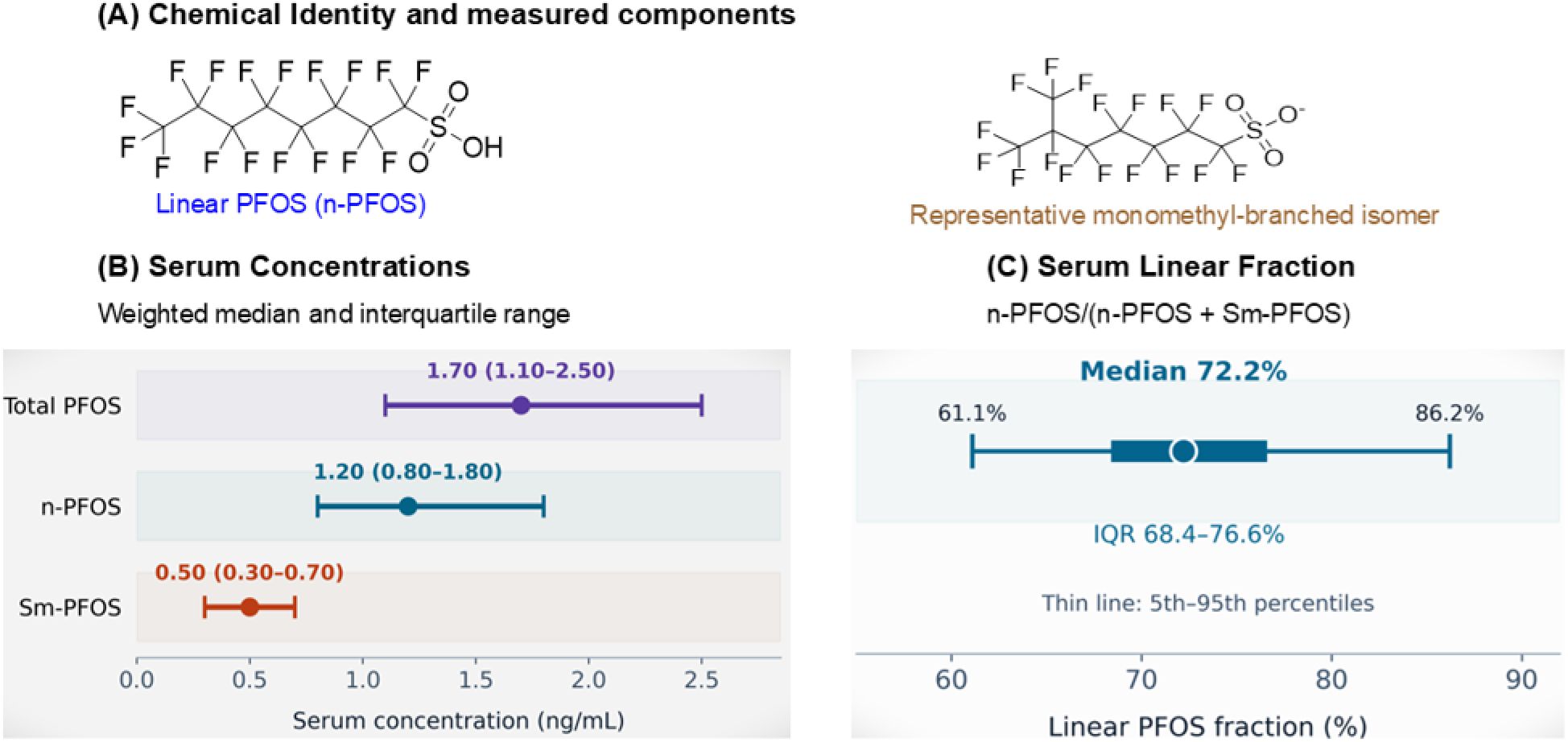
Chemical structures and serum PFOS distributions. (A) Condensed structures of linear PFOS and one representative monomethyl-branched isomer, shown as sulfonate anions. The 6-methyl structure is an example within the pooled Sm-PFOS measurement, not the structure of the entire pool. CDC identifies the 3-, 4-, 5-, and 6-methylheptane sulfonate isomers within Sm-PFOS; their individual proportions are not available in this analysis. (B) Points are weighted medians and bars are interquartile ranges in the 169-person PFOS–PHQ-9 cohort. (C) The linear fraction is n-PFOS/(n-PFOS + Sm-PFOS). The thin line spans the 5th–95th percentiles, the thick segment the interquartile range, and the point the median. These are serum measurements, not the composition of a commercial PFOS formulation. Quantiles were calculated separately, so the ratio of component medians need not equal the median individual ratio. No participant-level observations are reconstructed.

### 2.3 Depressive symptom outcomes

The PHQ-9 was administered through the audio computer-assisted interview system at the mobile examination center in this cycle.^3^ Each symptom item, DPQ010 through DPQ090, was scored from 0 to 3. The primary outcome was their sum, ranging from 0 to 27. Refused responses, “don’t know” responses, and missing values were treated as missing; all nine valid item responses were required. Item 9 was retained in the total score and was not analyzed as a separate suicidality outcome.

The secondary outcome was PHQ-9 ≥10. This threshold indicates at least moderate symptom severity for screening purposes, rather than a confirmed depressive disorder.^2^ A sensitivity outcome subtracted the sleep item (DPQ030) from the total score, yielding a range of 0–24. This modified score is not the validated PHQ-8, which excludes item 9, and no diagnostic threshold was assigned to it.

### 2.4 Covariates

Core covariates were age, sex, race and Hispanic origin, family income-to-poverty ratio, body mass index (BMI), and serum cotinine. Age, income-to-poverty ratio, and BMI were continuous. Sex used the NHANES male and female categories. RIDRETH3 was represented by its six categories; adjustment for this variable addressed social and population patterning rather than presumed biological differences in PFOS toxicity. Income-to-poverty ratio was top-coded at 5.00. BMI was obtained from the measured examination variable BMXBMI.

Serum cotinine (LBXCOT), a biomarker of recent nicotine exposure, entered as log₂(cotinine). Values below its 0.010 ng/mL LOD retained the NHANES-assigned fill value.^19^ Cotinine does not fully capture lifetime tobacco exposure or distinguish all sources of nicotine. Education was not included because the adult attainment variable is released for ages 20 years and older and would omit 18–19-year-olds. Sleep duration was not part of the core adjustment set because its causal position is uncertain and sleep symptoms contribute directly to the PHQ-9 outcome.

### 2.5 Survey regression and statistical inference

Analyses used the PFAS-specific 2-year subsample weight WTSPF2YR, supplied in PFAS_L, together with the masked stratum and primary sampling unit variables SDMVSTRA and SDMVPSU from DEMO_L.^9,18^ Variance estimates used Taylor linearization. Descriptive results are survey-weighted unless specified otherwise; participant counts and detection proportions are unweighted. Skewed concentrations are summarized by medians, interquartile ranges, and geometric means.

The primary model was survey-weighted linear regression of the continuous PHQ-9 score on both PFOS metrics and the core covariates:

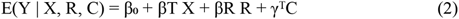

Here, Y is the PHQ-9 total, X = log₂(T), R = ln(L/B), and C is the covariate vector. The linear model estimates adjusted differences in mean symptom score under a linear conditional-mean specification. It does not assume that the symptom scale is a clinical diagnosis. Single-metric models were also fitted to show how estimates changed when concentration and composition were considered separately. A joint Wald test evaluated the two exposure terms together. Survey-weighted Pearson correlations described the relation between metrics; the reported pairwise variance inflation factor assessed their mutual collinearity, rather than collinearity with the complete covariate set. The coefficients do not provide a formal test that concentration has a greater association than composition because the exposure contrasts differ. Such comparisons also depend on their joint covariance and observed exposure support.

To make the scale of the estimates interpretable, we additionally rescaled the reported joint-model coefficients and their confidence limits to two fixed reference contrasts: total PFOS from 1.10 to 2.50 ng/mL, and linear fraction from 68.4% to 76.6%. These are the reported 25th and 75th percentiles in the 169-person descriptive cohort, not newly estimated percentiles of the adjusted sample. The multipliers are log₂(2.50/1.10) and log₂[(0.766/0.234)/(0.684/0.316)], respectively. These approximate contrasts use rounded summary values, retain the original model assumptions, and do not propagate uncertainty in the reference percentiles or represent additional model fitting (**Table S3**).

Survey-weighted logistic regression evaluated PHQ-9 ≥10 using the corresponding exposure specifications. Because the outcome was measured cross-sectionally, exponentiated coefficients are prevalence odds ratios, not incidence rate ratios or risk ratios. Estimates are reported with 95% confidence intervals and two-sided P values. The concentration and composition terms of the primary model were not adjusted for multiplicity; the confidence intervals are marginal rather than simultaneous.

### 2.6 Sensitivity analyses and missing data

Sensitivity analyses replaced the PHQ-9 total with the score omitting the sleep item, replaced the log-ratio with percent linear PFOS (per 10 percentage points), and restricted the joint model to participants with both components at or above their LODs. Sequential adjustment in the same 147 participants distinguished changes associated with covariate inclusion from changes associated with the complete-case sample. PFHxS, PFNA, and n-PFOA were evaluated separately as secondary exposures, each on the log₂ scale with core adjustment. Benjamini– Hochberg correction was applied across these three adjusted comparator tests.

Core analyses used complete cases. Among the 169 participants with complete PFOS and PHQ-9 measurements, 20 lacked income-to-poverty ratio and two lacked BMI; cotinine was complete. No multiple imputation was performed. Complete-case analysis is an analytic choice rather than a requirement of laboratory subsampling or PHQ-9 scoring, and its potential for selection bias is considered in the Discussion. Statistical significance was assessed at 0.05, but interpretation emphasized effect estimates, uncertainty, and sensitivity to model specification.

## 3 Results

### 3.1 Participant characteristics and PFOS distributions

The weighted mean age of the 169 participants was 21.2 years, and 56.5% were female (Table 1). The weighted median PHQ-9 score was 5 (interquartile range [IQR], 2-9). Forty-seven participants had PHQ-9 ≥10, corresponding to a weighted prevalence of 22.6%. In the 147 participants included in core-adjusted models, 42 had PHQ-9 ≥10 (weighted prevalence, 23.8%). These percentages describe positive screens within the respective analytic populations.

**Table 1.** Characteristics of participants aged 18-25 years with PFOS measurements and complete PHQ-9 scores in NHANES 2021-2023.

| Characteristic | n or count | Weighted estimate |
| --- | --- | --- |
| Age, years, mean (SE) | 169 | 21.2 (0.30) |
| Female, % | 94 | 56.5 |
| Mexican American, % | 22 | 10.2 |
| Other Hispanic, % | 22 | 12.5 |
| Non-Hispanic White, % | 88 | 54.5 |
| Non-Hispanic Black, % | 12 | 6.6 |
| Non-Hispanic Asian, % | 14 | 11.4 |
| Other or multiracial, % | 11 | 4.7 |
| Income-to-poverty ratio, median (IQR) | 149 | 1.91 (0.91–3.04) |
| BMI, kg/m<sup>2</sup>, mean (SE) | 167 | 26.4 (0.37) |
| Serum cotinine, ng/mL, median (IQR) | 169 | 0.021 (0.007–0.842) |
| PHQ-9 total, median (IQR) | 169 | 5 (2–9) |
| PHQ-9 $\geq 10$, % | 47 | 22.6 |
| n-PFOS ≥ LOD, unweighted % | 168/169 | 99.4 |
| n-PFOS, ng/mL, median (IQR) | 169 | 1.20 (0.80–1.80) |
| n-PFOS, ng/mL, geometric mean (95% CI) | 169 | 1.22 (1.13–1.31) |
| Sm-PFOS ≥ LOD, unweighted % | 166/169 | 98.2 |
| Sm-PFOS, ng/mL, median (IQR) | 169 | 0.50 (0.30–0.70) |
| Sm-PFOS, ng/mL, geometric mean (95% CI) | 169 | 0.44 (0.40–0.48) |
| ΣPFOS, ng/mL, median (IQR) | 169 | 1.70 (1.10–2.50) |
| ΣPFOS, ng/mL, geometric mean (95% CI) | 169 | 1.68 (1.57–1.80) |
| Linear PFOS fraction, %, median (IQR) | 169 | 72.2 (68.4–76.6) |
| IB_PFOS (ln ratio), median (IQR) | 169 | 0.96 (0.77–1.19) |
| log<sub>2</sub> linear-to-branched ratio, median (IQR) | 169 | 1.38 (1.12–1.71) |

The PFOS–PHQ-9 cohort includes 169 participants; core-adjusted models include 147. For categorical variables, n is the unweighted category count; for continuous variables, n is the available sample size. Detection counts and percentages are unweighted. Other estimates are survey-weighted. IQR, interquartile range; SE, standard error; CI, confidence interval. Total measured PFOS is n-PFOS + Sm-PFOS. The PFOS LOD is 0.10 ng/mL. IB\_PFOS denotes the natural-log ratio $\ln(\text{n-PFOS}/\text{Sm-PFOS})$ .

Linear PFOS was detected in 168 of 169 participants (99.4%), and Sm-PFOS in 166 (98.2%). Median total measured PFOS was 1.70 ng/mL (IQR, 1.10–2.50), with a geometric mean of 1.68 ng/mL (95% CI, 1.57-1.80). Median n-PFOS and Sm-PFOS concentrations were 1.20 and 0.50 ng/mL, respectively. The median linear fraction was 72.2% (IQR, 68.4-76.6%; 5^th^-95th percentiles, 61.1-86.2%; Figure 2). The median natural-log ratio was 0.96, corresponding to an individual linear-to-branched ratio of approximately 2.60.

The two component concentrations were positively correlated on the log₂ scale (weighted r = 0.787). In contrast, log₂(total PFOS) and isomer composition were weakly correlated (r = 0.215). In the complete-case sample, the corresponding correlation was 0.262 and the pairwise variance inflation factor was 1.07. Among all 194 age-eligible participants with valid PFAS data, the median linear fraction was 72.7% and the concentration–composition correlation was 0.251. Thus, the reported distributions did not suggest severe collinearity between the two exposure metrics, although weak correlation does not establish statistical or biological independence. The linear fraction varied by 8.2 percentage points across its IQR, whereas total PFOS varied 2.27-fold. These distributions show that participants can differ in composition without proportionate differences in total concentration; they do not identify distinct sources or independent toxic effects.

### 3.2 Associations with continuous depressive symptom burden

In the single-metric unadjusted model, each doubling of total PFOS was associated with a -0.92-point difference in PHQ-9 (95% CI, -1.71 to -0.12; P = 0.026). After core adjustment, the estimate was -0.45 points (95% CI, -1.44 to 0.54; P = 0.352). The single-metric composition estimate was 0.02 points per doubling of the ratio before adjustment (95% CI, -1.78 to 1.82) and 0.01 points after adjustment (95% CI, -1.97 to 2.00; Table 2).

**Table 2.** Associations of PFOS magnitude, isomer composition, and comparator PFAS with continuous PHQ-9 score.

| Exposure | Model | n | $\beta$ (95% CI) | P | q |
| --- | --- | --- | --- | --- | --- |
| ΣPFOS | Unadjusted | 169 | -0.92 (-1.71, -0.12) | 0.026 | — |
| ΣPFOS | Core-adjusted | 147 | -0.45 (-1.44, 0.54) | 0.352 | — |
| Isomer ratio | Unadjusted | 169 | 0.02 (-1.78, 1.82) | 0.983 | — |
| Isomer ratio | Core-adjusted | 147 | 0.01 (-1.97, 2.00) | 0.988 | — |
| ΣPFOS | Joint, unadjusted | 169 | -0.97 (-1.79, -0.15) | 0.024 | — |
| Isomer ratio | Joint, unadjusted | 169 | 0.32 (-1.56, 2.21) | 0.718 | — |
| ΣPFOS | Joint, core-adjusted | 147 | -0.49 (-1.65, 0.68) | 0.387 | — |
| Isomer ratio | Joint, core-adjusted | 147 | 0.21 (-1.94, 2.36) | 0.838 | — |
| PFHxS | Core-adjusted | 147 | -0.47 (-1.38, 0.44) | 0.291 | 0.371 |
| PFNA | Core-adjusted | 147 | -0.41 (-1.35, 0.53) | 0.371 | 0.371 |
| n-PFOA | Core-adjusted | 147 | -1.07 (-2.30, 0.16) | 0.085 | 0.255 |

$\beta$ is the PHQ-9 point difference per doubling of concentration or, for the isomer-ratio rows, per doubling of n-PFOS/Sm-PFOS. Joint models contain both $\log_2$ (total PFOS) and $\ln$ (n-PFOS/Sm-PFOS); other rows are single-metric models. Core adjustment includes age, sex, six categories of race and Hispanic origin, income-to-poverty ratio, BMI, and $\log_2$ serum cotinine. q values apply only to the three adjusted comparator tests. All models use WTSPF2YR, SDMVSTRA, and SDMVPSU. Values are rounded as reported.

In the primary joint model, the adjusted difference was -0.49 PHQ-9 points per doubling of total PFOS (95% CI, -1.65 to 0.68; P = 0.387) and 0.21 points per doubling of the linear-to-branched ratio (95% CI, -1.94 to 2.36; P = 0.838; Figure 3). The joint Wald test was F(2,15) = 0.46 (P = 0.638). These intervals include the null but also allow associations in either direction; they do not demonstrate equivalence of isomer compositions.

**Figure 3.**
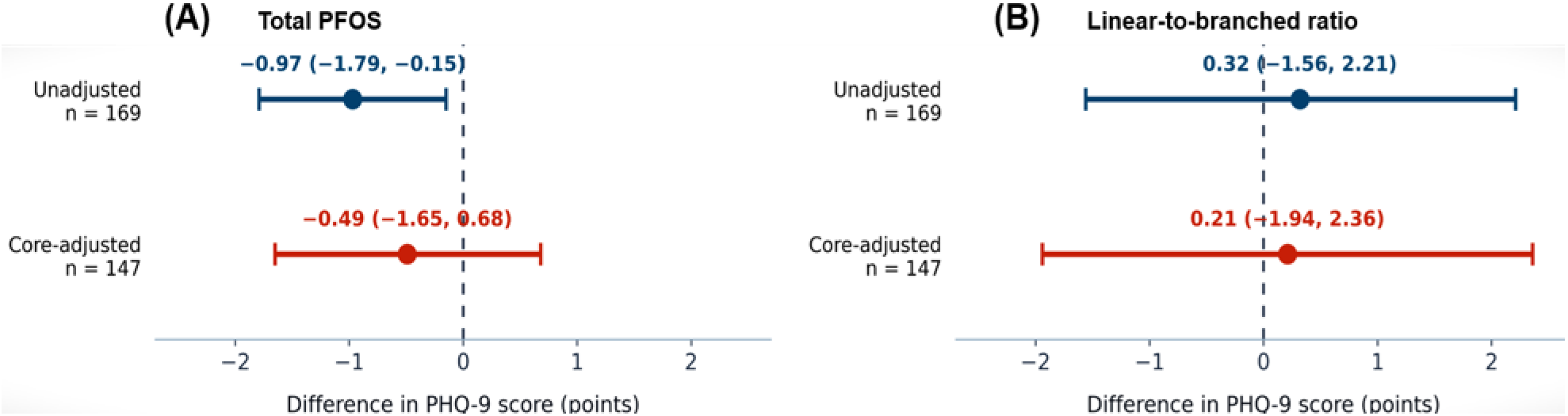
Joint associations of PFOS concentration and isomer composition with continuous PHQ-9 score. Both panels show terms from the same joint model at each adjustment level. (A) Difference per doubling of total PFOS at fixed composition. (B) Difference per doubling of n-PFOS/Sm-PFOS at fixed total PFOS. Points are coefficients and bars are 95% confidence intervals. Unadjusted models include 169 participants; core-adjusted models include 147 and adjust for age, sex, race and Hispanic origin, income-to-poverty ratio, BMI, and log₂ serum cotinine.

Across the reference total-PFOS contrast of 1.10 to 2.50 ng/mL, the joint model implied a −0.58-point PHQ-9 difference (95% CI, -1.95 to 0.81) at fixed composition. Across a linear-fraction contrast from 68.4% to 76.6%, it implied a 0.13-point difference (-1.16 to 1.41) at fixed total PFOS (**Table S3**). These algebraic translations place the estimates on the observed exposure scale. Their intervals overlap substantially, and their opposite point-estimate signs do not establish opposing biological effects.

The same-sample sequential analysis supported a role for adjustment in the attenuation of the continuous-score estimate. Among the 147 complete cases, the unadjusted total-PFOS coefficient was -0.90 (95% CI, -1.61 to -0.20). It changed to -0.63 after adjustment for age, sex, and race and Hispanic origin, to -0.42 after adding income-to-poverty ratio and BMI, and to −0.45 after adding cotinine (**Figure S1**). These changes show sensitivity to the included covariates; they do not isolate a causal contribution of any individual covariate.

### 3.3 Screening outcome and sensitivity analyses

For PHQ-9 ≥10, the single-metric odds ratio per doubling of total PFOS was 0.57 before adjustment (95% CI, 0.38-0.84; P = 0.008) and 0.59 after core adjustment (95% CI, 0.33-1.05; P = 0.070). Unlike the continuous-score coefficient, this point estimate changed little; the adjusted interval became wider and crossed the null. In the joint adjusted model, the composition odds ratio was 0.96 per doubling of the isomer ratio (95% CI, 0.27-3.44; P = 0.945).

Omission of the sleep item yielded an unadjusted total-PFOS coefficient of -0.76 (95% CI, -1.50 to -0.02) and a core-adjusted coefficient of -0.36 (95% CI, -1.28 to 0.56). The joint adjusted composition estimate for that outcome was -0.01 (95% CI, -2.07 to 2.05). With composition expressed as percent linear PFOS, the adjusted joint estimate was -0.02 points per 10 percentage points (95% CI, -1.61 to 1.57; **Table 3**).

**Table 3.** Screening outcome and sensitivity analyses.

| Outcome and model | Exposure contrast | n | Estimate (95% CI) | P |
| --- | --- | --- | --- | --- |
| Without sleep, unadjusted | Total PFOS ×2 | 169 | $\beta$ -0.76 (-1.50 to -0.02) | 0.045 |
| Without sleep, core-adjusted | Total PFOS ×2 | 147 | $\beta$ -0.36 (-1.28 to 0.56) | 0.417 |
| Without sleep, joint core-adjusted | Isomer ratio ×2 | 147 | $\beta$ -0.01 (-2.07 to 2.05) | 0.994 |
| PHQ-9 $\geq 10$, unadjusted | Total PFOS ×2 | 169 | OR 0.57 (0.38–0.84) | 0.008 |
| PHQ-9 $\geq 10$, core-adjusted | Total PFOS ×2 | 147 | OR 0.59 (0.33–1.05) | 0.070 |
| PHQ-9 $\geq 10$, joint core-adjusted | Isomer ratio ×2 | 147 | OR 0.96 (0.27–3.44) | 0.945 |
| PHQ-9 total, joint core-adjusted | Linear fraction +10 percentage points | 147 | $\beta$ -0.02 (-1.61 to 1.57) | 0.980 |
| PHQ-9 total, joint core-adjusted, both detected | Total PFOS ×2 | 144 | $\beta$ 0.02 (-0.98 to 1.01) | 0.974 |
| PHQ-9 total, joint core-adjusted, both detected | Isomer ratio ×2 | 144 | $\beta$ -0.98 (-2.34 to 0.39) | 0.148 |

Restriction to 144 complete-case participants with both components detected changed the joint total-PFOS coefficient from -0.49 to 0.02 (95% CI, -0.98 to 1.01) and the composition coefficient from 0.21 to -0.98 (95% CI, -2.34 to 0.39). All intervals included the null, but the shifts in direction and magnitude indicate that conclusions about numerical stability require caution. This restriction alters both the sample and the influence of substituted values, so the difference cannot be attributed to LOD substitution alone.

### 3.4 Secondary PFAS comparisons

PFHxS, PFNA, and n-PFOA were detected in approximately 96–99% of the cohort and were positively correlated with total PFOS on the log₂ scale (r = 0.51–0.65; **Figure S2**). The unadjusted n-PFOA coefficient was −1.28 PHQ-9 points per doubling (95% CI, −2.02 to −0.54). After core adjustment, it was −1.07 (95% CI, −2.30 to 0.16; P = 0.085; q = 0.255). Adjusted PFHxS and PFNA estimates also included the null (both q = 0.371; **Table 2**). The n-PFOA estimate was therefore already inconclusive before multiplicity correction. These secondary comparisons do not establish that inverse associations are specific to PFOS or to a particular PFAS functional group.

## 4 Discussion

The principal finding is that this small cross-sectional sample does not resolve whether serum PFOS composition is associated with depressive symptom burden independently of total concentration. The two exposure metrics were weakly correlated, making their joint inclusion feasible, but the adjusted intervals remained broad. A composition difference spanning the reported IQR corresponded to only 0.13 PHQ-9 points at the point estimate, yet its interval extended from -1.16 to 1.41. Consequently, the analysis provides neither evidence of a reliably different symptom burden across compositions nor an equivalence result. The chemical rationale for retaining isomer information is stronger than the statistical evidence for a composition association in this dataset.

Total PFOS and composition answer complementary questions. The concentration term compares higher and lower amounts of the measured mixture while holding its relative makeup constant. The composition term compares mixtures with different linear shares at the same total. A positive composition coefficient would be compatible with a higher linear share being associated with more symptoms, but would simultaneously represent a lower branched share. It could not assign causation to either component. Conversely, a near-zero coefficient could reflect similar biological effects, counteracting kinetic and toxicodynamic differences, an inadequate proxy for target-tissue dose, or insufficient precision. These explanations cannot be distinguished from a nonsignificant coefficient.

The model also does not establish that total PFOS has a greater impact than isomer composition. The reported -0.49 and 0.21 coefficients refer to different exposure changes, and both are uncertain. Rescaling to reference IQRs improves interpretability without converting them into comparable causal effects. Because Sm-PFOS combines several structures, two people with the same total PFOS and linear fraction could still have different distributions within the branched pool. The present two-component analysis cannot detect that additional chemical heterogeneity.

The exposure sequence begins with entry into the body, principally through ingestion of contaminated water or food, with contaminated dust and some occupational settings providing additional routes. The relative contribution varies by source and population; it was not measured here.^4^ After absorption, PFOS circulates in association with serum proteins and exchanges with tissues. The measured serum concentration combines bound and unbound material, whereas tissue delivery depends on binding, transport, tissue affinity, and clearance. Figure 4 separates this exposure and disposition sequence from the hypothesized downstream pathways to symptoms.

**Figure 4.**
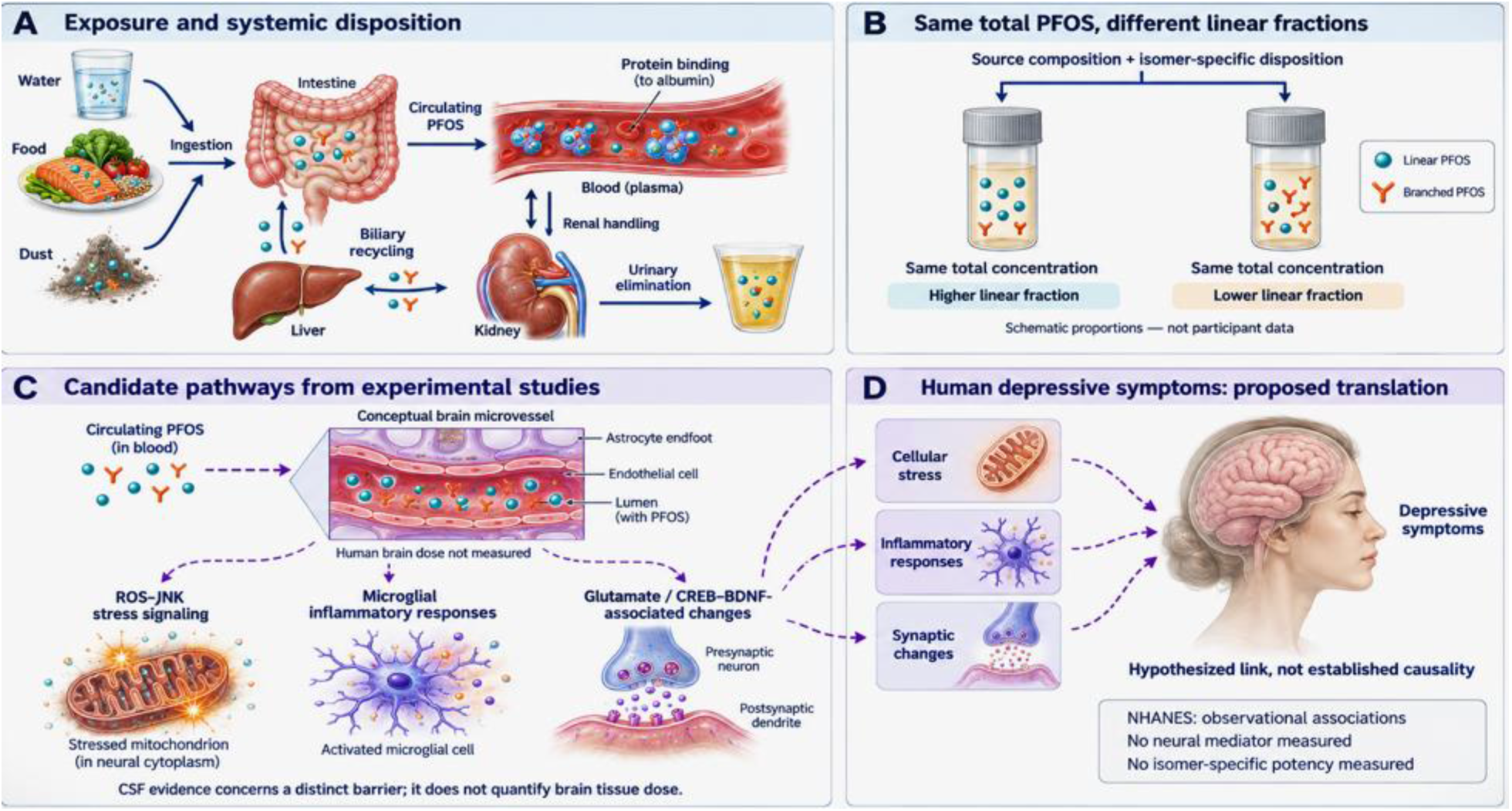
Exposure, disposition, and hypothesized pathways linking PFOS to depressive symptoms. (A) Ingestion of contaminated water, food, and dust can contribute to systemic exposure. Protein binding, renal handling, and biliary recycling influence circulating PFOS. (B) The same total measured concentration can have different linear fractions; source composition and isomer-specific disposition influence this ratio. Marker proportions are schematic and are not participant data. (C) Experimental studies support candidate ROS–JNK stress signaling, microglial inflammatory responses, and glutamate/CREB–BDNF-associated synaptic changes. The brain microvessel is a conceptual illustration; human cerebrospinal-fluid findings cited in the text concern a distinct barrier and do not directly measure brain tissue dose. (D) A link from these cellular processes to human depressive symptoms remains hypothetical. Solid arrows summarize exposure and disposition evidence; dashed arrows indicate proposed translation to neural effects and symptoms, not causal relationships established by NHANES.

Elimination is slow and involves more than a single renal process. In a small human study with four paired serum-bile samples, Harada and colleagues estimated substantial biliary reabsorption of PFOS, supporting enterohepatic recycling as one contributor to persistence.^20^ Human urinary measurements likewise support isomer-dependent renal handling.^13^ These findings help explain why a serum sample is not a direct measure of daily intake. They also create potential confounding pathways: kidney function, protein concentrations, and physiological elimination processes may alter the biomarker even when external exposure is similar.

The temporal scale matters for depression research. A persistent serum pool may reflect exposures accumulated over years, whereas PHQ-9 refers to the preceding 2 weeks. A cross-sectional association could therefore miss a vulnerable developmental window, a delayed effect, or an effect on persistent rather than recent symptoms. The current data cannot distinguish whether exposure preceded symptom onset. Changes in diet or other behaviors associated with symptoms could also modify subsequent exposure, producing reverse temporal ordering.

Branching provides a plausible basis for altered distribution, but the direction of a human neurological effect cannot be read directly from structure. In an isolated-albumin experiment, linear PFOS had a dissociation constant of 8 × 10⁻⁸ M, compared with approximately 8 × 10⁻⁵ to 4 × 10⁻⁴ M for the branched isomers studied.^11^ Tighter binding can support retention in the circulating compartment while reducing the unbound fraction under some conditions. These processes can have opposing implications for tissue delivery. The binding constants alone cannot specify brain exposure in an individual, because protein concentrations, competing ligands, transporter activity, and tissue clearance are not incorporated.

Human elimination data argue against treating all branched PFOS as one uniformly fast-clearing species. In airport employees followed after contaminated drinking water was replaced, Xu and colleagues estimated a linear-PFOS half-life of 2.93 years, compared with 1.05-1.26 years for measured branched isomers; 17 participants contributed monthly follow-up over 5 months.^14^ In contrast, Li and colleagues followed 114 Ronneby residents with up to ten samples over 2014-2018 and estimated 2.73 years for linear PFOS, 2.67 years for the 2/6-methyl group, and 3.43 years for the 3/4/5-methyl group.^15^ The studies differ in exposure history, observation period, isomer grouping, and model assumptions. Their results establish kinetic heterogeneity, not a universal ordering of linear versus branched persistence.

Barrier transfer is also context dependent. Greater branched-isomer transfer across the placenta in 20 matched maternal-cord pairs cannot be assumed to imply greater entry into the adult brain.^12^ Neonatal cerebrospinal-fluid measurements found that linear isomers were more frequently detectable than branched forms, but detection patterns depend on abundance, analytical sensitivity, developmental barrier properties, and partitioning.^21^ They do not establish a potency ranking in young adults. The mechanistic question is therefore whether a given serum composition changes target-tissue exposure or cellular response, not simply whether one structure is more abundant in blood.

This distinction yields competing, testable explanations for the same high linear fraction. It may mark a linear-enriched source mixture, preferential retention after exposure declines, or altered distribution between blood and tissues. A composition association might therefore arise without an intrinsic toxicity difference. Conversely, different intrinsic potencies could be obscured if the isomers produce different unbound or brain concentrations. The conceptual model in the Supporting Information makes these unmeasured quantities explicit; no kinetic or potency parameters were estimated from NHANES.

Entry into the central nervous system is possible, but serum PFOS cannot be equated with brain dose. In 223 matched serum-cerebrospinal-fluid pairs from hospital inpatients, Wang and colleagues reported mean PFOS concentrations of 6.8 ng/mL in serum and 0.028 ng/mL in cerebrospinal fluid. PFAS partitioning was associated with an albumin-based barrier-permeability index.^22^ Cerebrospinal fluid is not brain parenchyma, the blood-CSF barrier is anatomically distinct from the blood-brain barrier, and these clinical samples do not quantify penetration in healthy young adults. The observation supports restricted, variable access rather than free equilibration or proof that PFOS disrupts the barrier.

One candidate pathway is oxidative stress coupled to mitochondrial stress signaling. In SH-SY5Y cells, Sun and colleagues linked PFOS exposure to reactive oxygen species (ROS), JNK activation, mitochondrial and nuclear localization of phosphorylated JNK, and increased apoptotic signaling. Antioxidant treatment or JNK inhibition reduced the cellular response.^6^ These intervention experiments support a role for the ROS–JNK pathway in that cellular model. They do not demonstrate that neuronal apoptosis occurs at the serum concentrations in this cohort or that it mediates human depression. More subtle changes in neuronal function remain hypotheses requiring exposure-relevant testing.

A second candidate pathway involves microglial state and inflammatory signaling. Cheng and colleagues exposed cultured microglia to 25 or 50 μM PFOS for 24 hours and observed inflammatory transcriptional changes, impaired cellular functions, and altered H3K27ac profiles, some persisting after withdrawal.^7^ These findings provide a route by which PFOS could affect the cellular environment supporting synapses. They do not show that the same response occurs in human brain microglia at background exposure. For scale, the cohort median of 1.70 ng/mL is approximately 0.0034 μM using the PFOS-anion molecular mass.^23^ The experimental nominal concentrations are thus about 7,300–14,700 times that total-serum concentration. This comparison is descriptive, not a risk conversion: culture medium, protein binding, duration, and intracellular dose differ markedly from human exposure.

A third pathway connects glutamate regulation to synaptic plasticity. An and colleagues observed depressive-like behaviors in PFOS-exposed male mice, identified 105 dose-responsive differentially expressed hippocampal proteins, and reported increased glutamate with reduced glutamine and tryptophan. The study also found reduced CREB-BDNF signaling and lower PSD-95 and synaptophysin.^8^ Together, these observations support a hypothesis that disturbed excitatory signaling and impaired synaptic maintenance contribute to behavioral changes in that model. Mouse behavioral assays do not diagnose major depression, however, and these findings do not establish a linear-versus-branched potency difference.

The pathways in **Figure 4** should therefore be read as a sequence of hypotheses linking internal exposure to altered neural function and, potentially, symptoms. They are not mediators demonstrated in this study. No brain PFOS, unbound isomer concentrations, oxidative markers, cytokines, BDNF, or neural-function measures were available in the reported analysis. Moreover, evidence about a PFOS preparation cannot automatically be assigned to purified n-PFOS or any individual component of Sm-PFOS. Neither the direction nor the magnitude of an isomer-specific depression effect is established by the cited experiments.

The inverse concentration estimate for continuous PHQ-9 weakened after adjustment. In the same 147 complete cases, its magnitude decreased from 0.90 to 0.45 points per doubling, approximately 50%. The change therefore cannot be explained entirely by switching from the descriptive cohort to complete cases. It is compatible with confounding by the adjusted characteristics, but coefficient change does not identify a causal role for any one covariate. BMI and nicotine exposure, for example, may participate in several exposure–behavior–health relationships. The model remains dependent on its adjustment assumptions.

The screening outcome presents a different pattern. Its single-metric concentration odds ratio changed little, from 0.57 to 0.59, while the interval widened from 0.38–0.84 to 0.33–1.05. The crossing of P = 0.05 therefore primarily reflects precision rather than disappearance of the point estimate. Dichotomization also discards variation within PHQ-9 categories, and odds ratios are noncollapsible across adjustment sets. With only 42 positive screens in the adjusted sample, a model containing both exposures and multiple covariate parameters is vulnerable to imprecision. Neither outcome supports describing PFOS as protective.

External exposure patterns offer plausible noncausal explanations. For example, food sources contributing PFOS may also correlate with nutrients, income, or other determinants of mental health; source composition could additionally correlate with the isomer ratio. This is a confounding hypothesis, not a finding about diet in these participants. Symptom-related changes in eating, smoking, or health behaviors could also change exposure. The mixed directions reported in earlier NHANES studies reinforce the need for caution, especially because some studies use overlapping survey cycles and are not independent replications.^16,17^

The detection-restricted analysis is particularly informative about uncertainty. Excluding just three of 147 participants, approximately 2%, shifted the joint concentration estimate by +0.51 PHQ-9 points per doubling and the composition estimate by −1.19 points. The latter changed sign. Repeated confidence intervals spanning zero should therefore not be described as robust evidence of no association. Because low branched concentrations enter a denominator, relatively small absolute measurement changes can have substantial effects on the ratio. Exclusion also changes the weighted sample; observation and survey-cluster influence must be evaluated before attributing the difference specifically to LOD substitution.

The principal limitations are precision, selection, and exposure interpretation. The adjusted analysis contains 147 participants and a limited number of independent survey units. The 22 covariate-related exclusions represent 13.0% of the 169-person cohort; outcome completeness had already reduced the 194 participants with valid PFAS measurements to 169. Standard laboratory subsample weights do not necessarily correct selection from these additional exclusions. Similar median composition across the larger and smaller samples does not establish comparable symptom–exposure relationships. The cross-sectional design, single serum measurement, pooled branched component, and short symptom-assessment window further constrain causal interpretation.

Influence and missing-data sensitivity remain unresolved. The three detection-related exclusions warrant examination of weights, leverage, and leave-one-PSU-out estimates, alongside sensitivity to plausible values below detection. Missing-covariate analyses should compare complete cases with participants retained through design-aware multiple imputation under stated assumptions. Such analyses would address specific vulnerabilities identified here; they were not performed in this study.

Exposure-response shape and adjusted absolute probabilities would also improve interpretation if supported by the data. Parsimonious nonlinear terms could assess whether a linear coefficient conceals curvature, while marginal standardization could express screening associations as prevalence differences at specified concentrations and compositions. Predicted surfaces should be restricted to combinations actually supported by the sample, with uncertainty and data density shown. Extensive subgroup analyses, unrestricted interactions, or highly parameterized mixture methods would not overcome the limited sample and may amplify instability.

A stronger epidemiologic test would use repeated exposure and symptom measurements or carefully harmonized additional NHANES cycles. Pooling requires appropriate weights and attention to assay definitions and the change to audio computer-assisted PHQ-9 administration in 2021-2023.^3,10^ Mechanistic follow-up should compare purified linear and individual branched isomers at matched unbound or measured intracellular concentrations, with defined isomer mixtures, physiological protein conditions, and exposure durations relevant to the hypothesis. Measuring trans-barrier transport before ROS, microglial, and synaptic responses would help distinguish kinetic differences from intrinsic potency. These are testable extensions, not conclusions that can be supplied by the present cross-sectional associations.

## 5. Conclusions

Total serum PFOS concentration and its linear-to-branched composition describe different features of internal exposure. In young adults in NHANES 2021-2023, joint models did not establish an independent composition association with depressive symptoms, and wide intervals and sensitivity to a few observations limit inference. Experimental studies support possible oxidative, inflammatory, and synaptic pathways, but serum composition reflects disposition as well as intake and cannot rank the depression-related toxicity of PFOS isomers. The findings motivate reproducible, larger, and temporally informative analyses that connect isomer-resolved exposure to biologically relevant internal dose.

## Data Availability

The public NHANES data files used for this analysis are available from the National Center for Health Statistics at https://wwwn.cdc.gov/nchs/nhanes/continuousnhanes/default.aspx?Cycle=2021-2023. Table S1 lists the files and variables needed to identify the reported exposures, outcomes, covariates, and survey design.

## Supporting information

The Supporting Information provides the variable dictionary, exposure and coefficient transformations, derived reference contrasts, a conceptual exposure-to-response model, missing-data and detection notes, sequential model estimates, and PFAS correlations.

