## Supporting Information for "Serum PFOS Concentration and Isomer Composition in Relation to Depressive Symptoms in Young Adults"

### S1 Data sources and variable definitions

Public-use data from NHANES August 2021–August 2023 were linked by respondent sequence number (SEQN). The primary analysis used the five files listed in Table S1. The PFAS-specific weight WTSPF2YR is supplied in PFAS_L; the masked survey design variables are supplied in DEMO_L. A positive PFAS subsample weight was required.

**Table S1. Files and variables used in the reported analyses**

| **File** | **Variables** | **Use** |
| --- | --- | --- |
| DEMO_L | SEQN; RIDAGEYR; RIAGENDR; RIDRETH3; INDFMPIR; SDMVSTRA; SDMVPSU | Linkage; age; sex; race and Hispanic origin; income-to-poverty ratio; survey strata and primary sampling units |
| PFAS_L | SEQN; WTSPF2YR; LBXNFOS; LBXMFOS; LBDNFOSL; LBDMFOSL | PFAS subsample weight; linear and summed monomethyl-branched PFOS; detection flags |
| PFAS_L | LBXPFHS; LBXPFNA; LBXNFOA; LBDPFHSL; LBDPFNAL; LBDNFOAL | Secondary PFHxS, PFNA, and n-PFOA concentrations and detection flags |
| DPQ_L | SEQN; DPQ010; DPQ020; DPQ030; DPQ040; DPQ050; DPQ060; DPQ070; DPQ080; DPQ090 | Nine PHQ-9 symptom items; DPQ030 is the sleep item |
| BMX_L | SEQN; BMXBMI | Measured body mass index |
| COT_L | SEQN; LBXCOT; LBDCOTLC | Serum cotinine and detection flag; cotinine LOD 0.010 ng/mL |

All files are from NHANES August 2021–August 2023. Concentration values below detection retain the public-file fill values. Flag value 0 denotes at or above detection; 1 denotes below detection. Sources: the corresponding NCHS file codebooks cited in the main manuscript.

### S2. Magnitude and composition parameterization

For each participant, let L = LBXNFOS and B = LBXMFOS, with both concentrations in ng/mL. The operational total, linear fraction, natural-log ratio, and log₂ ratio are related as follows:

T = L + B; f = L/T; R = ln(L/B); Z = log₂(L/B)	(S1)

R = ln[f/(1 − f)]; Z = R/ln(2); f = exp(R)/[1 + exp(R)]	(S2)

The pair (T, R) retains the information in the two measured concentrations when both are positive. The inverse transformation is:

L = T × exp(R)/[1 + exp(R)]; B = T/[1 + exp(R)]	(S3)

Consequently, concentration and composition are distinct coordinates of the same measurements rather than independent exposure sources. The positive NHANES fill values permit the transformations for nondetections, but do not eliminate measurement uncertainty near the LOD.

#### S2.1 Regression coefficient scaling

The main model uses X = log₂(T) and R = ln(L/B). If the fitted composition coefficient and confidence limits on the natural-log scale are bR, lR, and uR, the corresponding values per doubling of L/B are:

bdouble = bR ln(2); CIdouble = [lR ln(2), uR ln(2)]	(S4)

For a logistic model with natural-log-ratio coefficient aR, the odds ratio per doubling is exp[aR ln(2)], and its confidence limits are obtained by applying the same transformation to the coefficient limits. If Z = log₂(L/B) is fitted directly, its coefficient is already expressed per doubling and must not be rescaled again.

The original notation IB_PFOS corresponds to R. All isomer-ratio estimates in Tables 2 and 3 and Figure 3 are expressed per doubling of L/B, regardless of the natural-log scale used in fitting. The percent-linear sensitivity analysis uses 100f, with its coefficient expressed per 10 percentage points. Because the log-ratio and percent-linear transformations differ, linear models on the two scales impose different functional forms.

#### S2.2 Meaning of a fixed total composition contrast

At fixed T, doubling the linear-to-branched ratio changes f to 2f/(1 + f). At f = 0.722, this gives approximately 0.839. This calculation illustrates the regression contrast and is not an observed longitudinal change or a causal intervention. Conversely, doubling T at fixed R doubles L and B together.

### S3. Outcomes detection and missing data

The PHQ-9 total is the sum of DPQ010 through DPQ090 after restricting each item to valid responses 0, 1, 2, or 3. Codes 7 and 9, and missing values, are excluded from scoring. Complete responses to all nine items are required. The score omitting sleep is PHQ-9 total minus DPQ030; it ranges from 0 to 24 and is not the validated PHQ-8. PHQ-9 ≥10 is a screening outcome.

The LOD is 0.10 ng/mL for n-PFOS and Sm-PFOS. LBDNFOSL and LBDMFOSL equal 0 for measurements at or above the LOD and 1 for measurements below it. The released concentration fields retain the NHANES fill value of LOD/√2 at the precision of the public file. Detection was reported in 168/169 participants for n-PFOS and 166/169 for Sm-PFOS. The paired-detection sensitivity analysis included 144 of the 147 complete cases. High overall detection does not rule out influence from the three excluded observations.

Core covariates were complete in 147 of the 169 participants with PFOS and PHQ-9 data. Twenty participants lacked income-to-poverty ratio and two lacked BMI; cotinine was complete. No multiple imputation was performed. Laboratory subsampling, missing biospecimens, incomplete outcomes, and missing covariates represent distinct stages of selection. The standard PFAS weight addresses the subsample design and does not, by itself, establish that additional complete-case exclusions are ignorable.

### S4. Model definitions and inference

Single-metric models contain either X or R. Joint models contain both. Core adjustment includes continuous age, the NHANES sex category, six categories of race and Hispanic origin, continuous income-to-poverty ratio, measured BMI, and log₂ serum cotinine. Models incorporate WTSPF2YR, SDMVSTRA, and SDMVPSU. Linear-regression estimates are differences in mean PHQ-9 points; logistic-regression estimates are prevalence odds ratios.

The primary exposure pair is evaluated without multiplicity adjustment. Secondary PFHxS, PFNA, and n-PFOA analyses each use a separate core-adjusted model. Their three P values constitute the Benjamini–Hochberg family: 0.291, 0.371, and 0.085, yielding q values of 0.371, 0.371, and 0.255, respectively. The correction is not applied to an unspecified collection of unreported models.

The reported pairwise variance inflation factor of 1.07 is consistent with 1/(1 − r²) for r = 0.262 in the complete-case sample. It describes collinearity between the two exposure metrics only and does not establish that the full design matrix is well conditioned. The correlation of 0.215 pertains to the larger 169-person cohort.

### S5. Additional results

Sequential adjustment among the same 147 participants is summarized in Table S2 and Figure S1. Maintaining the same sample distinguishes coefficient changes associated with covariate inclusion from changes caused by excluding incomplete cases. Figure S2 presents the pairwise PFAS correlations. Because total PFOS contains both isomer components, its strong correlations with each component partly reflect the shared arithmetic construction and should not be interpreted as independent confirmation of a common exposure source.

**Table S2. Sequential concentration estimates in the same complete-case sample**

| **Model** | **n** | **β per doubling of total PFOS** |
| --- | --- | --- |
| Unadjusted | 147 | −0.90 |
| Age, sex, race and Hispanic origin | 147 | −0.63 |
| Previous model plus income-to-poverty ratio and BMI | 147 | −0.42 |
| Previous model plus log₂ cotinine | 147 | −0.45 |
| Previous model plus isomer composition | 147 | −0.49 |

The outcome is continuous PHQ-9 score. The reported unadjusted 95% CI is −1.61 to −0.20; the core-adjusted CI is −1.44 to 0.54; the joint core-adjusted CI is −1.65 to 0.68. Intermediate confidence intervals are displayed in Figure S1. The sequence assesses sensitivity to adjustment and does not identify the causal contribution of individual covariates.

### S6. Derived exposure contrasts

The calculations below use rounded coefficients and confidence limits from the joint core-adjusted model in Table 2 and reference percentiles from the 169-person descriptive cohort. They are algebraic translations of that model, not newly fitted models, observed changes within participants, or estimates of adjusted-sample percentiles. Uncertainty in the chosen reference values is not propagated.

ΔY = bT log₂(T₂/T₁) + bd log₂[(f₂/(1 − f₂))/(f₁/(1 − f₁))]	(S5)

**Table S3. Approximate PHQ-9 contrasts derived from the joint core-adjusted model**

| **Reference contrast** | **Multiplier** | **PHQ-9 difference (95% CI)** |
| --- | --- | --- |
| Total PFOS 1.10 → 2.50 ng/mL at fixed composition | 1.1844 | −0.58 (−1.95 to 0.81) |
| Linear fraction 68.4% → 76.6% at fixed total PFOS | 0.5968 | 0.13 (−1.16 to 1.41) |

Reference percentiles are from the n = 169 descriptive cohort. Estimates rescale the n = 147 joint model and inherit its model uncertainty. These are fixed-reference contrasts, not newly fitted models.

Here bT is the coefficient per doubling of total PFOS and bd is the coefficient per doubling of L/B. For a contrast changing only one coordinate, multiply that coefficient and both confidence limits by the corresponding positive log₂ contrast. Total PFOS from 1.10 to 2.50 ng/mL gives a multiplier of 1.1844. Linear fraction from 0.684 to 0.766 gives a multiplier of 0.5968. These reference contrasts do not establish that all corresponding combinations of T, f, and covariates have adequate sample support.

For a contrast changing both coordinates, the joint coefficient covariance is required to calculate its uncertainty. If a = log₂(T₂/T₁) and c = log₂[(f₂/(1 − f₂))/(f₁/(1 − f₁))], then:

Var(ΔY) = a²Var(bT) + c²Var(bd) + 2ac Cov(bT, bd)	(S6)

Only single-coordinate reference contrasts are reported. Joint-change confidence intervals, formal contrasts between exposure coefficients, and adjusted probability surfaces would require the corresponding joint-model covariance and were not calculated. A joint-model concentration odds ratio cannot be reconstructed from the single-metric odds ratio.

### S7. Conceptual separation of disposition and biological response

The following equation is a conceptual accounting model to clarify interpretation, not a fitted dose–response model. Let f be the linear serum fraction and wj the fraction of branched isomer j within Sm-PFOS, with Σwj = 1. Let aL and aBj summarize the combination of tissue delivery and response per unit serum concentration under specified experimental conditions:

H = T [ f aL + (1 − f) Σj wj aBj ]	(S7)

H denotes a hypothetical biological perturbation and is not a PHQ-9 prediction. The coefficients may depend on unbound fraction, tissue transport, intracellular exposure, and intrinsic potency; they are not established constants. Under this simplified model, changing total concentration scales the mixture at fixed composition. Changing f at fixed T changes H in proportion to aL − Σj wj aBj. The sign is unknown without experimental data. Further, the same measured f can conceal different wj values. No wj, aL, aBj, or H values were measured or estimated in this study.

An exposure-source explanation predicts composition changes with source mixture even without altered intrinsic potency. A disposition explanation predicts associations of serum composition with albumin, kidney function, or time since exposure reduction. A toxicodynamic explanation requires different responses after matching unbound or target-cell dose. These explanations can coexist; the serum ratio alone cannot discriminate among them.

### S8. Experimental concentration comparison

Using the PFOS-anion molecular mass of 499.12 g/mol, 1.70 ng/mL equals 1.70 μg/L, or approximately 3.4 nmol/L (0.0034 μmol/L). The 25 and 50 μmol/L nominal culture concentrations in Cheng et al. are approximately 7,300 and 14,700 times this median total-serum concentration. These are arithmetic comparisons, not human-equivalent doses or safety margins. They do not account for culture-medium composition, free concentration, intracellular accumulation, exposure duration, or tissue distribution. References: main manuscript references 7 and 23.

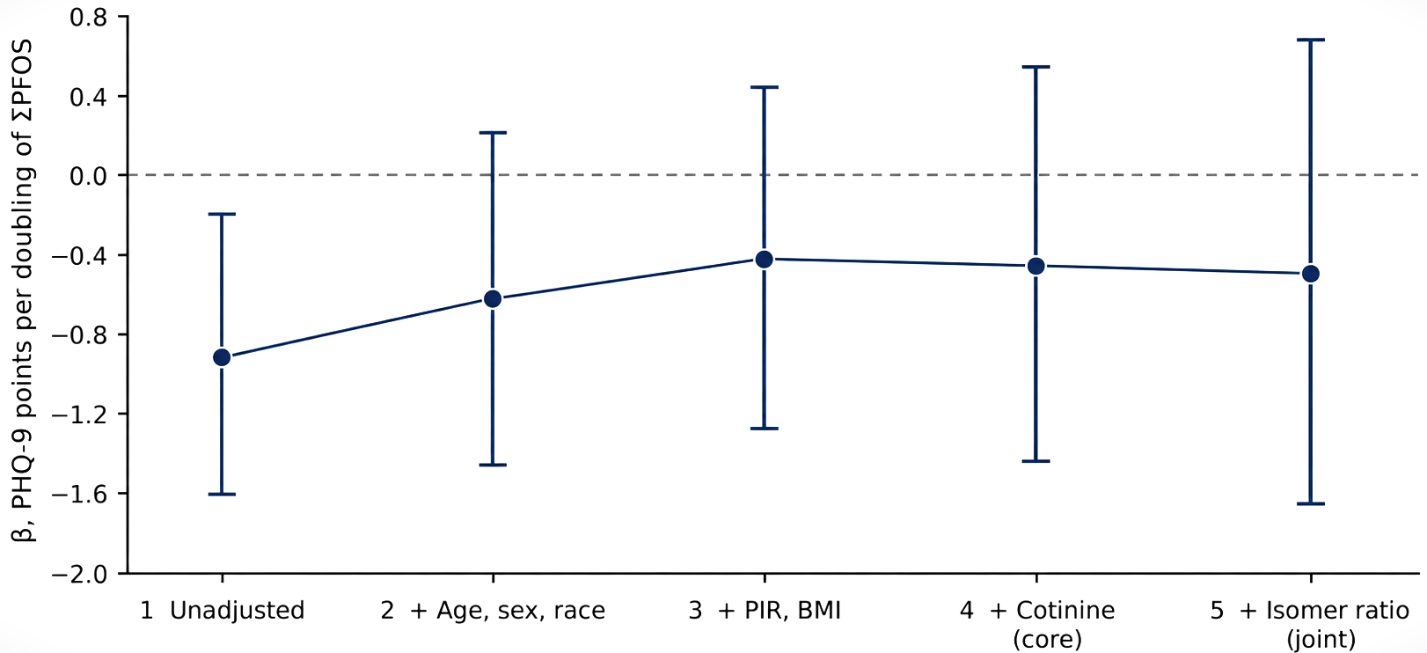

**Figure S1.** Sequential adjustment of the total-PFOS association with continuous PHQ-9 in the same 147 participants. Each point is the coefficient per doubling of total PFOS, and bars denote reported 95% confidence intervals. Models successively add age, sex, and race and Hispanic origin; income-to-poverty ratio (PIR) and BMI; log₂ cotinine; and the isomer-ratio term. The connecting line indicates model order rather than a time trajectory. Estimates incorporate the PFAS subsample weight and survey design. Coefficients are listed in Table S2.

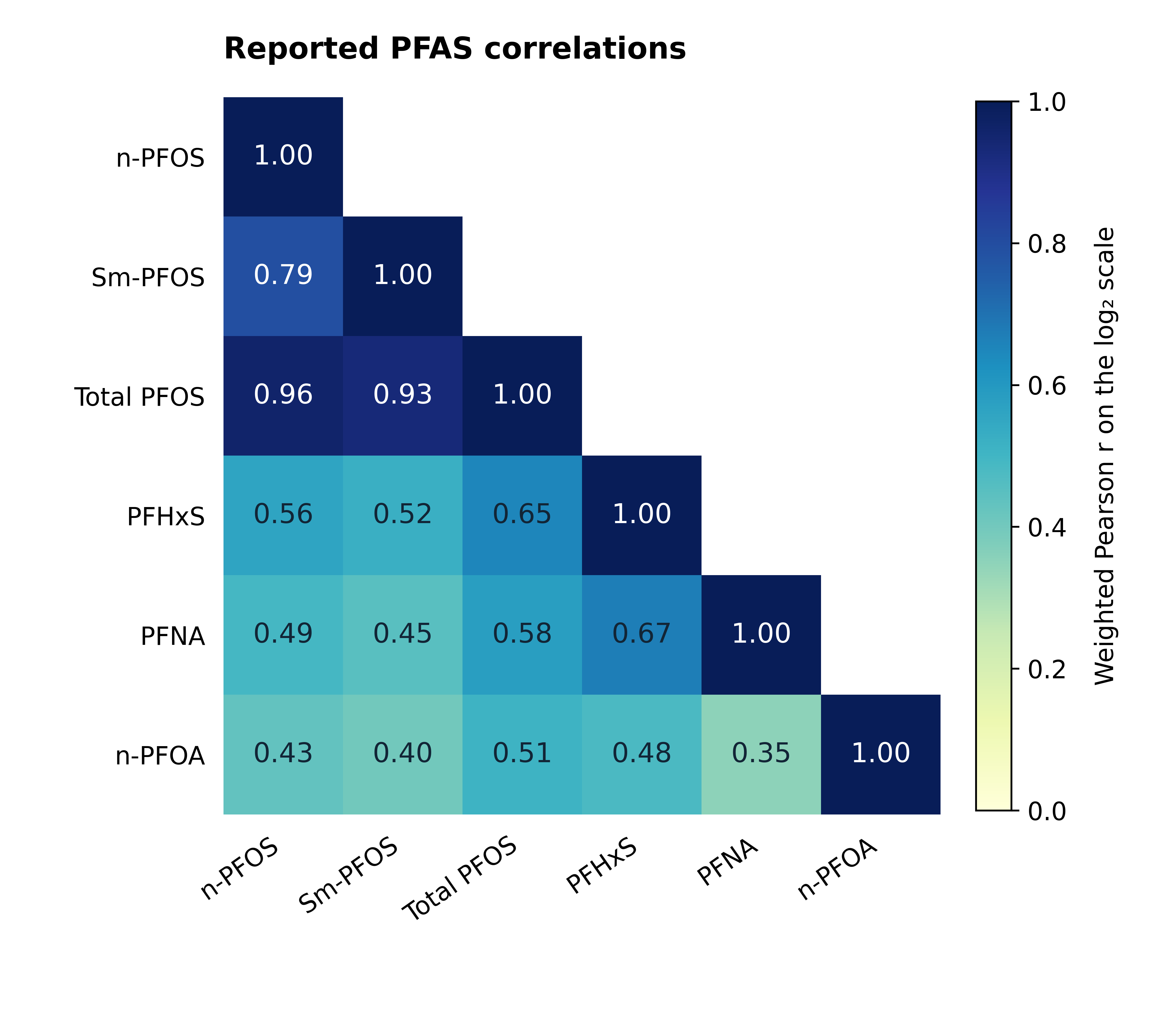

**Figure S2**. Survey-weighted pairwise Pearson correlations among log₂-transformed PFAS concentrations in the 169-person PFOS–PHQ-9 cohort. Total PFOS is the sum of n-PFOS and Sm-PFOS. Correlations of total PFOS with its components partly reflect this arithmetic relationship. The heatmap redraws the reported matrix rounded to two decimals; correlations were not recalculated for this illustration. The composition ratio is excluded because it is a relative metric rather than an additional concentration. Its correlation with log₂(total PFOS), reported separately in the main text, is 0.215. Concentration values below detection retain the NHANES-assigned fill values. Weights: WTSPF2YR.
